# Standards for reporting norms in the scientific literature and the development of free-access tools to apply them in practice

**DOI:** 10.1101/2024.11.12.24317147

**Authors:** Rok Blagus, Bojan Leskošek, Francisco B. Ortega, Grant R. Tomkinson, Gregor Jurak

## Abstract

Norm-referenced tests compare individuals to a group. While norms are often presented in tables and graphs, exact score evaluation relies on model parameters, often undisclosed. These models, like those from the R *gamlss* package, include individual data protected by law and consent, hindering full transparency. Thus, this paper proposes standards for publishing test norms that allow precise score evaluation while protecting participant privacy. We outline specific requirements for norms publications: a) the exact presentation of the fitted model that contains the estimates of all model parameters and other information required for exact evaluation; b) computer sharable fit of the model that does not contain any sensitive information and can be used by those with programming skills to evaluate scores; and c) a web-based application that can be used by those without programming skills to use the results of the fitted model. To facilitate publication and utilization of norms, we have developed and provided in this manuscript an open-source R package of tools for authors and users alike.

## 1. Introduction

In scientific work, measurement plays a crucial role in evaluating the characteristics of objects or events. Various tests—often referred to as measures, instruments, or scales—are employed to gather data, which can be classified as either criterion-referenced or norm-referenced^1^. Criterion-referenced tests measure characteristics against a fixed criterion to establish cut-offs (also called cut-points or thresholds), for example, adult body mass scores 17, 23, 28, and 33 kg/m2 are classified as underweight, normal, overweight, and obese, respectively, based on adult cut-offs of 18.5, 25, and 30 kg/m2^2^. Norm-referenced tests are designed to assess and compare an individual’s performance against a pre-established reference or source population (e.g., national, regional, or global population comparator). These tests are used across various fields including psychology, education, and healthcare to evaluate corresponding abilities or attributes^3^. For example, in psychology, norm-referenced tests like the Wechsler Intelligence Scale for Children (WISC) are used to assess intellectual abilities^4^. In education, standardized tests like the SAT (Scholastic Aptitude Test) and ACT (American College Test) help determine student placement and readiness for college^5^. Similarly, in healthcare, growth charts based on norms for height and mass are essential tools for pediatricians to monitor child development^6^. In sports, norm-referenced tests are often used for performance evaluation (e.g., fitness or skill-based profiling and monitoring)^7,8^, talent identification^9^, and to inform coaching strategies^10^.

In scientific literature, norms —also referred to as normative data or normative values— are typically tabulated and visualized. Tables often display the specific centile values (e.g., 1^st^, 10^th^, 25^th^, 50^th^) at various age intervals (e.g. 20-29y, 30-39y, etc.), while graphs display centile curves or bands^11^. These resources often include only select centiles and age intervals, making it difficult to determine exact centile ranks for specific ages^12^. This lack of detailed data needed to calculate the exact centiles and for exact ages is exacerbated when additive terms, such as P-splines, are required^13^. These transformations necessitate detailed model parameters, which are often not fully disclosed in published papers, limiting the ability of professionals and test-takers to apply the norms accurately^12^. Consequently, the effective use of norm-referenced tests in practice demands not only comprehensive normative data but also accessible computational tools to facilitate precise and efficient calculation.

Among the many tools used to generate models for normative evaluation of test-takers’ scores, probably the most widely used nowadays is the *gamlss* library in R that implements Generalized Additive Models for Location, Scale, and Shape (GAMLSS). For example, it has been used to create reference centile curves for measures of body size in pediatric studies, allowing for more accurate assessments of child growth patterns^14^, for fitness tests in monitoring the motor development of children^8^ and monitoring changes across the adult lifespan^15^. However, despite of widespread use of *gamlss* library in normative research, there’s a serious drawback: the authors are not allowed to publish the GAMLSS models as they contain all the individual values of variables used for generating the model, which are usually protected by law (e.g. the general data protection rule (GDPR) in the European Union (EU)) and/or study participant’s informed consent agreement. So even though *gamlss* library contains functions (e.g. *centile*.*pred*) to easily and precisely evaluate test-taker’s score, field professionals cannot use those functions as they don’t possess the normative model.

Accordingly, this paper recommends the standards for the publication of norms that will enable the exact normative evaluation of test-taker’s score while respecting the privacy of personal data used in norms construction. We also develop and describe the tools that will enable authors to efficiently publish the norms in accordance with the standard and enable end-users (practitioners, test-takers etc.) with or without programming skills to compute the exact normative scores. These tools include: a) a tool for generating a publication-ready, human-readable report in a typical setting (R GAMLLS model with several predictor variables, of which several are potentially modelled using P-splines); b) a tool for generating machine-readable object, which may be used by norms authors for publishing norms (e.g. as supplementary file to the norm’s paper) or by anyone to develop an app which will (by using accompanying methods) support exact norm calculations to the end-users with no programming experience; c) a tool which enables the authors of the norms with little programming experience to easily generate a web app which enables exact calculation of normative values to the end-user with no programming skills.

## 2. Construction of norms using GAMLSS

GAMLSS enables the modeling of four parameters of a distribution: *mu, sigma, nu*, and *tau*, related to location, variation, skewness, and kurtosis, respectively, as some functions of the explanatory variables. The distribution does not necessarily belong to the exponential family^16^. While some distributions are completely characterized by only some of the four parameters, e.g. the Gaussian distribution that only requires *mu* and *sigma*, mean and standard deviation, respectively, the distributions commonly used to construct norms, e.g. the Box-Cox Power Exponential distribution, depend on all four parameters^17^. An appropriate link function relating the distribution parameters to the explanatory variables needs to be chosen to ensure that the predictions for the modeled parameter are within the range of values that the parameter can take. E.g. for the Gaussian distribution the *sigma* parameter needs to be positive, hence the link function needs to be some monotonic function that takes a positive value as an argument and returns a real number. The *gamlss* R library offers reasonable default specifications of the link functions for each available distribution: e.g. for the Gaussian distribution the default link functions are the identity and the natural logarithm functions for *mu* and *sigma*, respectively.

When constructing the norms, it is common to include additive terms that enable modeling non-linear associations. These additive terms can be defined in numerous ways, regression splines are a commonly used approach (see Perpreroglou et al.^18^ for an overview of using splines to model non-linear associations with a focus on the R software). In norms construction, P-splines, a popular version of penalized regression splines^19^, are often used. P-splines try to overcome some issues of the other types of splines, e.g., the dependence on the number and position of the knots – the points within the data range where adjacent smooth functional pieces (usually low-order polynomials used to fit the data between two consecutive knots) join each other. P-splines generally use cubic B-spline basis with many equidistant knots and a penalty term which reduces the potential problem of overfitting due to many knots^20,21^. The amount of penalty is controlled by the smoothing parameter. When constructing the norms Generalized Akaike Information Criterion (GAIC) is commonly used to determine the optimal amount of smoothing^22^.

The fitted GAMLSS model can be used to construct the norms by estimating the parameters of the assumed distribution for the particular values of the explanatory variables and evaluating the distribution function (i.e. the probability that some variable does not exceed a particular value) or its inverse, the quantile function (more details are given in the Supplementary material).

## 3. Standards for the publication of norms

A paper that presents norms should contain the following information, in the form of supplementary material.

1. Details about the fitted model, in the form of a table and as a computer-readable object (e.g. as an R object), that contains the following information.
  1. The family used to model the outcome (e.g. the Box-Cox Power Exponential distribution).
  2. Link functions used to model each parameter of the distribution (e.g. the identity function, the natural logarithm function, the identity function and the natural logarithm function, for the *mu, sigma, nu*, and *tau* parameters of the Box-Cox Power Exponential distribution, respectively).
  3. The estimated linear coefficients for each parameter.
  4. If the model includes additive terms, the estimated penalized coefficients and further details needed to completely recreate the B-spline basis of the P-spline for the parameters where the additive terms are present.
2. A fully functioning web application enabling those without programming knowledge to use the published norms.

The tabulated information will enable the readers to better comprehend the underlying model and will make it easier to understand the differences between the published models. Researchers familiar with computer programming, e.g., R, will be able to use the computer-readable object for the exact evaluation of the test taker’s score. However, since this requires computer programming skills, a fully functioning web application enabling those without any programming knowledge to use the published norms should be designed and published. We developed the necessary tools to represent the fitted GAMLSS model according to the above standard. These tools are described in detail in the next section.

## 4. Tools for publishing the norms

The tools that can be used to publish the norms in accordance with the standards from the previous Section are available as the R package *gamlssReport* published on GitHub (rokblagus/gamlssReport). The R package is easy to install in R via *install_github*(“*rokblagus*/*gamlssReport*”).

There are two main functions:

1. the function *gamlssReport* extracts all the necessary information from the fitted GAMLSS and represents it in the Table format and as an R object;
2. the function *ShinyApp*.*gamlssReport* builds the web application.

Our implementation allows multiple additive terms modeled by using P-splines. The package also contains other functions (e.g. *centile*.*gamlssReport*), which can be used to evaluate the test-taker’s score by only requiring the output of the function *gamlssReport*.

## 5. An example

We illustrate how to use the package by providing an example. We provide all the necessary R code, appearing after the R> symbol, that is required to present the fitted model in accordance to the standard or to evaluate the test taker’s score within R. We assume that the users of the package can fit the GAMLSS model using the R package *gamlss*.

Our example is based on the FitBack dataset, where we focus on the standing long jump (SLJ) measurements for boys. This is one of the most popular muscular fitness tests in the world. FitBack dataset includes 1,383,773 test results of SLJ from 31 European countries^8^. Before fitting the model, the data were cleaned as described in Ortega et al.^8^ GAMLSS was then fitted in R (using R version 3.6.3^23^) assuming Box-Cox t distribution, modeling all four parameters of the distribution as a non-linear function of age, using P-splines, optimizing the smoothing parameter using the Schwarz Bayesian criterion (SBC)^24^. Power transformation was used for *age* before including it in the model (i.e. a variable *nage=age*^*1/2*^ was included in the model).

After fitting the model, stored in R as an R object *fit*, all the necessary information required to evaluate test-taker’s score, is obtained by using the function *gamlssReport* using the object created by the GAMLSS package as the argument:

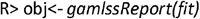

The function *print* using the object generated by the function *gamlssReport* then displays the model in the table format:

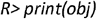

For our FitBack example, the printed object is represented in Figure 1.

**Figure 1:**
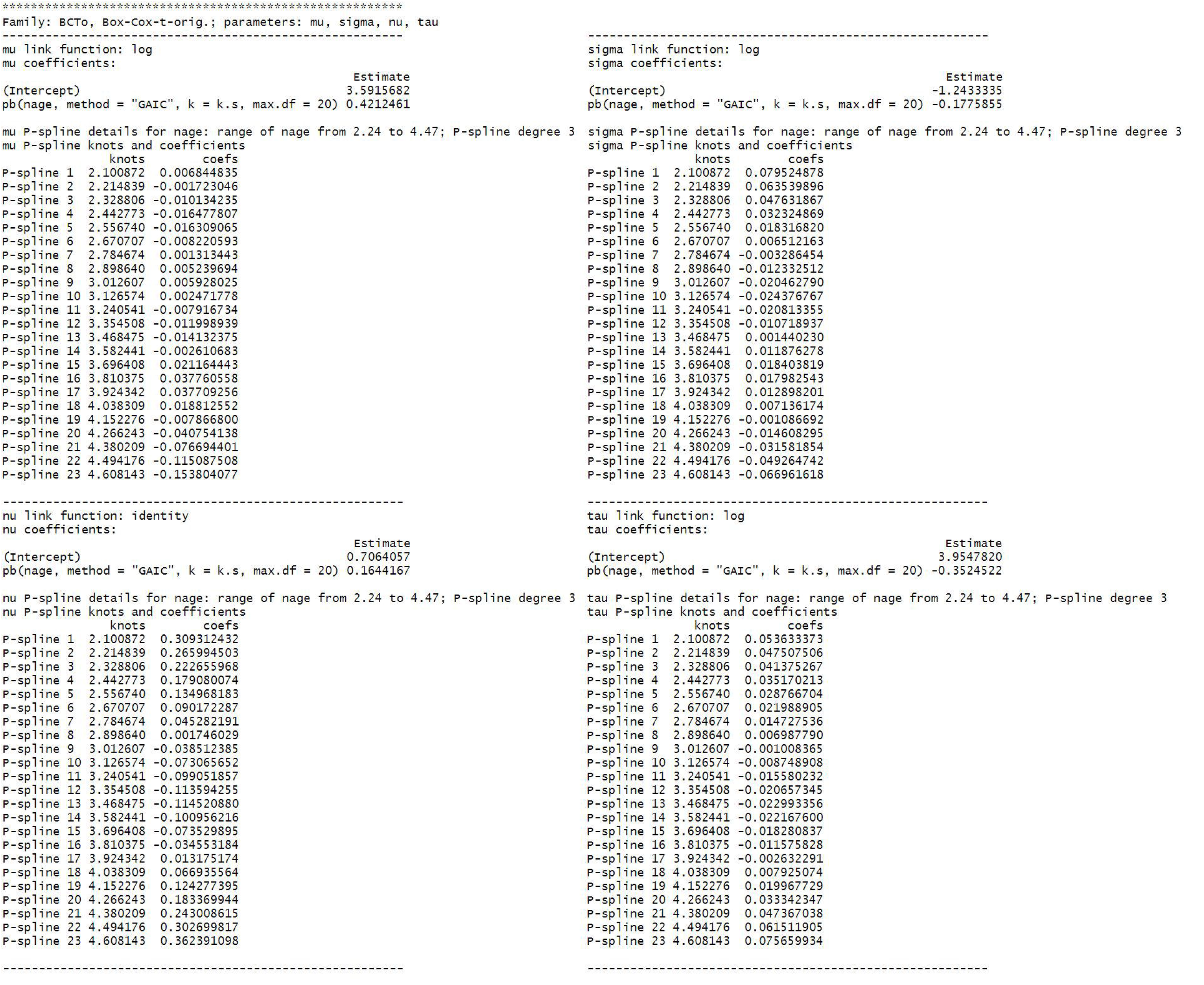
Fitted GAMLSS model for the FitBack data; Standing Long Jump test (cm) for boys.

Figure 1 first reports the assumed distribution and the list of its parameters. Then there are four blocks of results, one for each parameter. In each block, the link function used for modeling a certain parameter (e.g. log for the *mu* parameter), the linear coefficients (e.g. 3.60 for the *intercept* and 0.42 for the transformed age - *nage* for the *mu* parameter), the range of the variable used in P-splines (e.g. 2.24 to 4.47 for *nage* for the *mu* parameter) and the degree of the polynomial used when forming the spline (e.g. 3 for the *mu* parameter), the knots (e.g. 2.10,…,4.61 for the 23 knots for the *mu* parameter) and their respective penalized coefficients (e.g. 0.01,…,-0.15 for the 20+3=23 penalized coefficients for the *mu* parameter) are reported. While the penalized coefficients cannot be directly interpreted, they are vital for estimating the parameters of the fitted distribution which is required to evaluate the test-taker’s score as illustrated in detail in the Supplementary material.

To calculate the centile for a 10-year-old boy whose SLJ was 140 cm, we can use the function *centile*.*gamlssReport*:

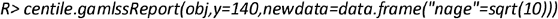

The function *centile*.*gamlssReport* takes the object obtained by the function *gamlssReport* as the first argument, score(s) for which the centile(s) is (are) to be calculated as the second argument, and a data frame containing the values of all the variables for which the centiles are to be calculated (in our example we only need to supply the value *10*^*(1/2)*^ for the variable *nage*, the sole variable in our model) as the final argument. In our example, the function *centile*.*gamlssReport* returns the value 54.7, i.e. the boy’s score corresponds to 54.7th centile. It is also possible to obtain the estimated parameters of the assumed distribution, that were required to calculate the centile, using the function *predict* (see Supplementary material for more details).

To see which score corresponds to e.g., the 90th centile for 10-year-old boys we can use the function *score*.*gamlssReport*:

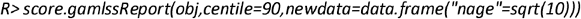

The function *score*.*gamlssReport* has similar arguments as the function *centile*.*gamlssReport*, but instead of the score it takes the argument *centile* which represents the centile(s) for which to calculate the score(s). In our example the function returns 166.4, meaning that the 90th centile for 10-year old boys on SLJ is 166.4 cm.

Another useful function in our package is the *plot* function, which plots the centile curves. This function extends the function *centiles* from the *gamlss* R library by allowing multiple explanatory variables when fitting the model (in the function centiles only one explanatory variable is supported). When there are more explanatory variables in the model, one variable for which the centile curves will be displayed needs to be chosen (this is set via the argument *xname* in the function *plot*) while the other variables are set to some value (e.g. to their respective mean or mode), which is controlled by the argument *newdata*. Using this function for our SLJ example:

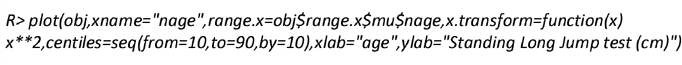

yields the plot presented in Figure 2. The function enables transforming the x-axis so that the centile curves are represented on the original scale when using the power transformation (in the above example we show the centiles as a function of *age* and not *nage*).

**Figure 2:**
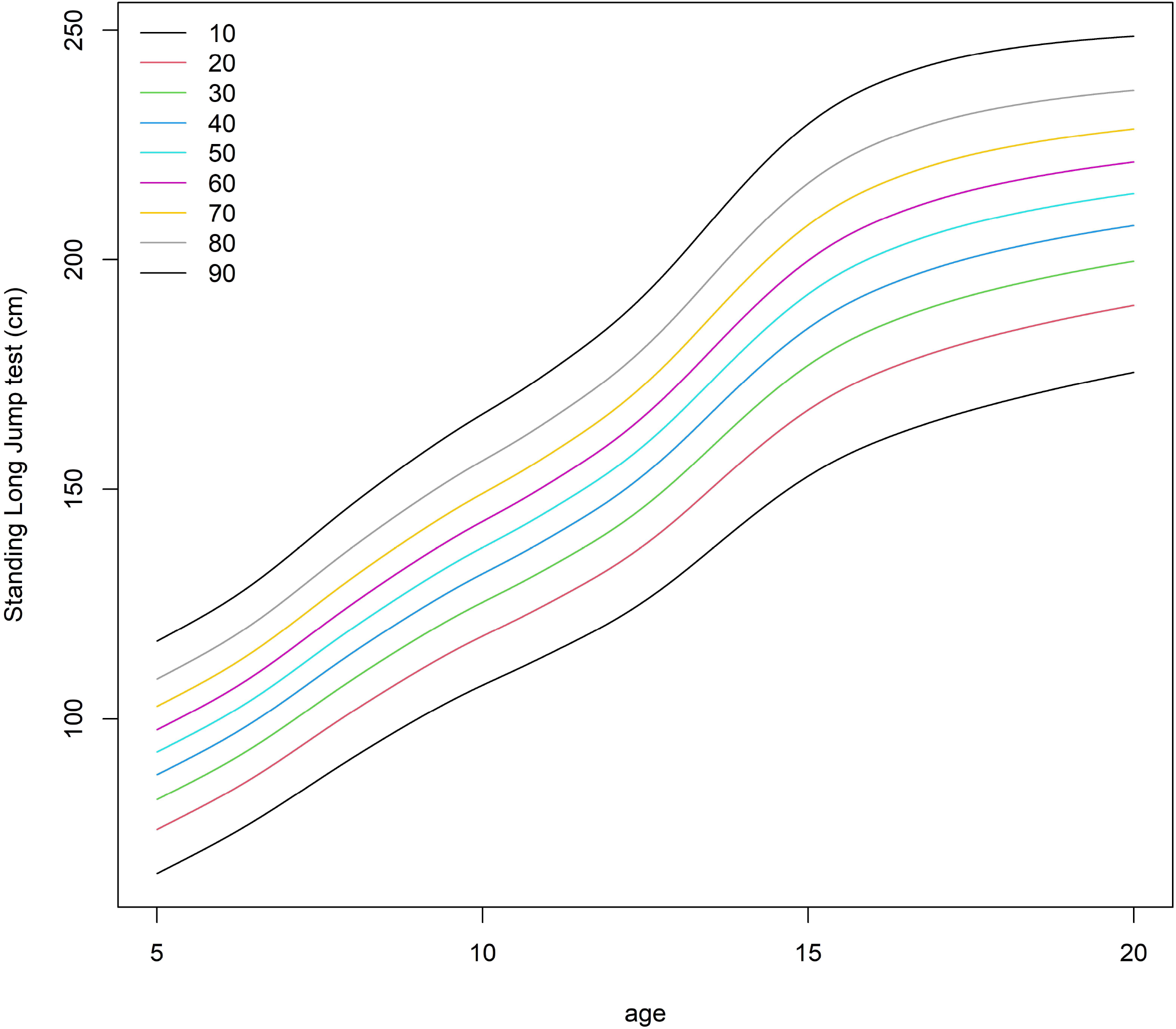
The centile curves produced by the function plot, using only the object generated by the function *gamlssReport*.

To produce a fully functional web app, the authors of the norms can use the function *ShinyApp*.*gamlssReport*. If the authors want the app to be publicly accessible, the app should be uploaded to a server that supports Shiny, for example, https://www.shinyapps.io/.

Using the function *ShinyApp*.*gamlssReport* for our example produces an app which is displayed in Figure 3. This app can be used to calculate the centile by entering the *age* and the *score* (in our example we set the age to 10 and the score to 140 cm in which case the app evaluates the centile) or the score by entering the *age* and the *centile* (in which case the app would report the score at the given *centile* bellow the plot). It is not difficult to use the functions available in our R package and the R package *Shiny* to produce more complex web-based model summaries such as the one published on https://leska.shinyapps.io/FitBack/ where we summarize the norms for all the tests from the ALPHA-fit battery for both genders that were published in Ortega et al.^8^

**Figure 3:**
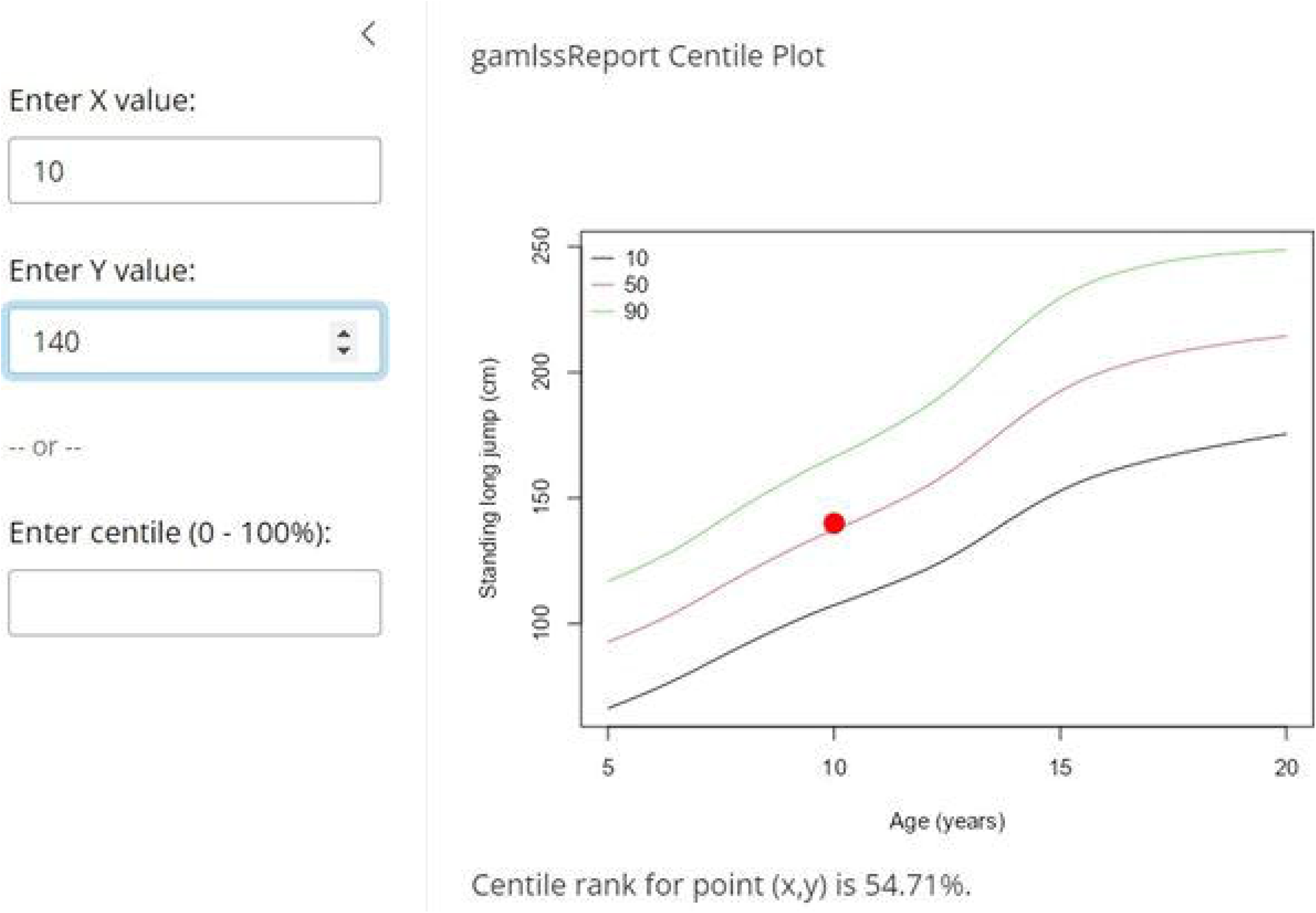
Shinny app for the FitBack data; Standing Long Jump test (cm), boys. The example shown is for evaluating the score of a 10-year-old boy with a score of 140 cm (red point).

## 6. Conclusions

We recommend a standard for publishing normative data that enables the exact interpretation (exact centile for exact age) of an individual observation evaluated, without sharing any potentially sensitive information that is protected by law (e.g., GDPR in EU) and/or study participant’s informed consent agreement. Further, it enables a comparison of different published models, by showing all the details of the fitted model. In this manuscript, we have developed, described, and made freely available the necessary tools to publish the norms in accordance with this standard, using R language for statistical computing. We illustrated the tools using the FitBack dataset as an example.

## Data availability statement

The original FitBack dataset analyzed during the current study is not publicly available due to violating confidentiality but is available upon a reasonable request. The R object *obj* produced by the function *gamlssReport* is published on GitHub (rokblagus/*gamlssReport*/obj.Rdata).

## Acknowledgments

This work was supported by the Slovenian Research and Innovation Agency - ARIS (Methodology for data analysis in medical sciences, P3-0154; Bio-psycho-social research program, P5-0142).

## Author Contributions

RB – conceptualization, data curation, formal analysis, investigation, methodology, software, validation, visualization, writing – original draft preparation, writing – review and editing.

BL – conceptualization, data curation, formal analysis, investigation, methodology, software, validation, visualization, writing – original draft preparation, writing – review and editing.

FBO – data curation, writing – review and editing.

GJ – data curation, funding acquisition, writing – review and editing.

GRT – writing – review and editing.

All authors have read and approved the final version of the manuscript, and agree with the order of presentation of the authors.

