## Supplementary file for "Standards for reporting norms in the scientific literature and the development of free-access tools to apply them in practice"

#### Supplementary material

Blagus, Rok^1,2^; Leskošek, Bojan^3^; Ortega, Francisco B.^4,5^; Tomkinson, Grant R.^6^; Jurak, Gregor^3^

^1^ Institute for Biostatistics and Medical Informatics, University of Ljubljana, Ljubljana, Slovenia

^2^ Faculty of Mathematics, Natural Sciences and Information Technologies, University of Primorska, Koper, Slovenia

^3^ Faculty of Sport, University of Ljubljana, Ljubljana, Slovenia

^4^ Department of Physical Education and Sports, Faculty of Sport Sciences, Sport and Health University Research Institute (iMUDS), University of Granada; and CIBEROBN Physiopathology of Obesity and Nutrition; Granada, Spain

^5^ Faculty of Sport and Health Sciences, University of Jyväskylä, Jyväskylä, Finland.

^6^ Alliance for Research in Exercise, Nutrition and Activity (ARENA), Allied Health and Human Performance, University of South Australia, Adelaide, SA, Australia

### Using the fitted model to evaluate the test taker’s score

The centile for some test score, say *y*, given the values of the explanatory variables, say *x*, is obtained by using *x* to estimate the parameters of the distribution (this entails using the inverse of the link function, e.g. when the link function is the natural logarithm function, its inverse, the exponential function is used to estimate the parameter). After obtaining the estimated parameters, the distribution is completely specified (at this *x*), and the centile is then easily obtained as a value of the distribution function at *y*. For example, let’s assume that the estimated mu and sigma for the normal distribution are 0 and 1, respectively, and that *y*=1.96. Then the centile is 0.975. The score for some centile is obtained similarly by using the quantile function (i.e., the inverse of the distribution function). For the previous example, the score for the 2.5^th^ centile would be, using the quantile function of the standard normal distribution, - 1.96.

### Application to the FitBack example

The results reported in Figure 1 in the main document can be used to evaluate the test taker’s score as follows. Assume that a 10-year-old boy’s score on the standing long jump (SLJ) was 140 cm.

First, we need to estimate each parameter of the assumed Box-Cox-t-distribution for *nage*=10^(1/2)^=3.16 (recall that a power transformation of age was used). To do this, we need to obtain 23 values for the B-spline basis for each of the 23 penalized coefficients (in our case these will be the same for all four parameters since the same spline is used for all parameters; the predictions will, however, differ since the penalized coefficients differ for each parameter), denote these values when presented as a vector with B. E.g., in R we can use the *splineDesign* function from the *splines* package (some care must be taken so that the function behaves as the gamlss’ function *pb*, therefore also the range for the variable used to represent the spline is included in the output), to obtain B=(0,0,0,0,0,0,0,0,0.05,0.58,0.36,0.01,0,0,0,0,0,0,0,0,0,0,0)^T.

In our R package we use the *gamlss*’ internal function *bbase* to form the spline basis, but using the function *splineDesign* would yield identical results. We can then estimate log(*mu*)=3.60+0.42*3.16+0*(0.01)+...+0*(-0.15)=4.92, from where we obtain *mu*=exp(4.92)=137.35. After a similar calculation, using different linear coefficients and penalized coefficients from their respective blocks from Figure 1 in the main document, we obtain *sigma*=0.16, *nu*=1.15, and *tau*=16.94. In our R package these predictions can be obtained with the function *predict* using the object created by the function *gamlssReport*:

*R> predict(obj, newdata=data.frame("nage"=sqrt(10)))*

The function *predict* takes two arguments: the object obtained by the function *gamlssReport* and *newdata* which is a data frame containing the values of all the variables needed to make predictions (in our case the model contains only the variable *nage)* for which the predictions are to be made (in our example *sqrt(10)*).

To get the centile, we then calculate the value of the distribution function of the Box-Cox t distribution using the estimated parameters for the score equal to 140 cm (in R this can be obtained using the function *pBCTo* from the *gamlss* package), which yields 0.547, i.e. the boy‘s score corresponds to 54.7th centile.

To see which score corresponds to e.g., 90th centile for 10-year-old boys, we evaluate the quantile function of the Box-Cox t distribution at 0.9, again using the estimated parameters (in R this can be done using the *gamlss*’ function *qBCTo*), yielding 166.4 cm.

### A technical note

The approach that we described for calculating the predicted values, in the presence of additive terms, differs slightly from the approach that is used in *gamlss*’ *predict* function. The *predict* function in the *gamlss* package works as follows. The predictions considering only the penalized coefficients for the additive term, separately for each additive term, are obtained for each observation that was used to fit the model. These are then used to perform cubic spline interpolation (in R this is e.g. available in the function *spline*) for the data points for which the predictions are to be calculated. The resulting values are then added to the predictions obtained from the linear coefficients, again using the inverse of the link function to finally obtain the estimated parameter. The two approaches yield the same results when calculating the predictions for the values that were used when fitting the model but can differ for the values not observed during the fitting, especially at the boundaries or when two sequential data points in the original dataset are far apart.

These differences can however mainly be attributed to the method that *gamlss* is using when doing the interpolation. In particular, the implementation in *gamlss* is using natural splines when doing interpolation (i.e. their approach yields the same result as when using the R function *spline* with the argument x set to the observed values of the variable that is used to form the spline basis, y set to the fitted value for the smooth term (i.e. the fitted values using the penalized coefficients), *xout* set to new values for which the interpolation is being performed and the argument method set to *natural*). If on the other hand, the argument method would be set to *fmm*, resulting in the spline of (Forsythe et al., 1979) (an exact cubic is fitted through the four points at each end of the data, and this is used to determine the end conditions), the results would be very similar to our approach that was described above. The differences between the two approaches will in general not be substantial, and while the approach based on natural splines might be more suitable when doing extrapolation (i.e. doing predictions outside the observed range), this is never done within the context of centile estimation. Furthermore, note that when using the approach that is implemented in the *gamlss* R package, all original datapoints of the variables used to form the splines have to be shared in order to use the model’s results, which might be problematic in some applications, as already discussed in the introduction. In our package we offer the possibility that the calculations are performed as in the *gamlss* package (setting the argument *extract.smooth* in the *gamlssReport* function to *TRUE*; the default is *FALSE*), yielding identical results also for the datapoints not observed when fitting the model. However, the exported R object in this case contains potentially sensitive information that the user might not want to share.
